# Effects of Maternal Fortified Balanced Energy-Protein Supplementation on the Mother-Infant Gut Microbiome: A Sub-Study of the MISAME-III Randomized Controlled Trial

**DOI:** 10.1101/2023.11.24.23298964

**Authors:** Lishi Deng, Steff Taelman, Matthew R. Olm, Laéticia Céline Toé, Eva Balini, Lionel Ouédraogo, Yuri Bastos-Moreira, Alemayehu Argaw, Kokeb Tesfamariam, Erica D. Sonnenburg, Moctar Ouédraogo, Rasmané Ganaba, Wim van Criekinge, Patrick Kolsteren, Michiel Stock, Carl Lachat, Justin L. Sonnenburg, Trenton Dailey-Chwalibóg

**Affiliations:** Department of Food Technology, Safety and Health, Faculty of Bioscience Engineering, Ghent University, Ghent, Belgium; BIOBIX, Department of Data Analysis and Mathematical Modelling, Ghent University, 9000 Ghent, Belgium; KERMIT, Department of Data Analysis and Mathematical Modelling, Ghent University, 9000 Ghent, Belgium; BioLizard nv, Ghent, Belgium; Department of Microbiology and Immunology, Stanford University School of Medicine, Stanford, CA, USA; Institut de Recherche en Sciences de la Santé (IRSS), Unité Nutrition et Maladies Métaboliques; Centre Muraz, Bobo-Dioulasso, Burkina Faso; Center of Excellence in Mycotoxicology and Public Health, MYTOXSOUTH® Coordination Unit, Faculty of Pharmaceutical Sciences, Ghent University, 9000 Ghent, Belgium; Agence de Formation de Recherche et d’Expertise en Santé pour l’Afrique (AFRICSanté), Bobo-Dioulasso, Burkina Faso; Chan Zuckerberg Biohub, San Francisco, CA, USA; Center for Human Microbiome Studies, Stanford University School of Medicine, Stanford, CA, USA

## Abstract

Biological pathways, including individual gut microbiome are potential barriers for maternal nutritional supplementation to improvement in infant growth. We evaluated the impact of balanced energy-protein (BEP) supplementation during pregnancy and the first six months of lactation on the composition and functionality of gut microbiome in mothers and their infants in rural Burkina Faso. Our findings reveal that BEP supplementation led to a significant increase in microbiome diversity during pregnancy. In the second trimester, there was a notable decrease in the abundance of an *Oscillospiraceae* species, while postpartum, the abundance of *Bacteroides fragilis* increased. We identified concerted enriched or depleted microbial pathways associated with BEP supplementation, including the phosphotransferase system, a critical mechanism for bacterial carbohydrates uptake, which exhibited enrichment in infants born to BEP-supplemented mothers. Despite these observations, the intricate biological connections with other omics necessitate further analysis to fully elucidate the underlying comprehensive biological pathways.

## Introduction

It estimated that 148.1 million children under the age of 5 – about 22% of the global population - experienced stunted growth in 2022, hindering their full developmental potential^1^. Stunted growth commonly begins before birth, so-called fetal growth restriction and accumulates during infancy until the age of 18 to 23 months; after 2 years of age, the growth impairment tends to be largely irreversible^2–5^. Maternal undernutrition in pregnancy is a major risk factor for poor child growth^2,5^. A growing body of research has investigated the effect of maternal nutritional supplementation on infant growth, such as lipid-based nutrient supplement (LNS) and balanced energy-protein (BEP), but the results have been mixed^6–10^. These findings suggest that the role of biological pathways needs to be considered.

From the first colonization of the neonate at birth through maternal microbiota, succession in bacterial taxa differ based on geographical area^11–13^, feeding practice^14,15^, dietary patterns, age, and health status^16,17^. A more mature gut microbiota at the first month may improve infant growth trajectories in the first year of life^18^. Inadequate maturation of the gut microbiome might contribute to the onset of child malnutrition, and both microbiome immaturity and malnutrition can be partially ameliorated following nutrition interventions^19,20^. Therefore, given its plastic nature, the individual gut microbiome appears to be a promising barrier to improvement in growth outcomes.

The evidence on the effect of maternal nutritional supplementation on infant gut microbiome is scarce. Two analyses based on randomized controlled trials (RCTs) in Malawi showed inconsistent results. The first study (*n* = 869) revealed a significant enhancement in alpha diversity among infants receiving LNS from 6 to 18 months^21^, while the second study (*n* = 631) demonstrated no statistically significant effects of LNS on the diversity and maturation of the infant gut microbiota in samples collected between 6 and 30 months of age^22^.

BEP supplements are a similar type of ready-to-use supplements that provide multiple micronutrients and, specifically, energy and protein in a balanced composition (<25% of total energy content from protein) to address maternal malnutrition during pregnancy and lactation. We previously reported that, in the MIcronutriments pour la SAnté de la Mere et de l’Enfant-III (MISAME-III) trial, BEP supplementation increase energy and macro- and micronutrient intakes among pregnant women and fills nutrient gaps without displacing food intake^23^. Moreover, we observed a modest effect of prenatal BEP supplementation on weight and length increments at birth, and on the prevalence of low birth weight^24^. The improvements in size at birth accrued from prenatal BEP supplementation were sustained in terms of linear growth at the age of 6 months^7^. However, the impact of providing pregnant and lactating women with BEP supplementation on gut microbiota has yet to be investigated.

The objective of the present study was to evaluate the efficacy of BEP supplementation during pregnancy and the first six months of lactation in influencing the gut microbiota in both mothers and infants, compared to the control group who received iron folic acid (IFA) tablets. This encompasses an examination of the maternal and infant metagenome in terms of microbial composition and functionality, including microbial metabolic pathways, a dimension that has not been explored in prior studies.

## Results

### Study population

A total of 731 fecal samples from 152 pregnant and lactating women and their infants (*n* = 71 in BEP group and *n* = 81 in IFA group) were collected and sequenced; these comprised 256 infant samples and 475 maternal samples. In the IFA group, infant samples were taken at two time points, 1-2 months (76 samples) and 5-6 months (56 samples) of life, and maternal samples were taken at four time points, inclusion (i.e., trimester 2, 20 samples), trimester 3 (78 samples), 1-2 months (76 samples) and 5-6 months (75 samples) postpartum (total IFA samples = 381). In the BEP group, 66 and 58 infant samples were taken at 1-2 months and 5-6 months of life, and 22, 71, 66 and 67 maternal samples were taken at inclusion, trimester 3, 1-2 months and 5-6 months postpartum (total BEP samples = 350) (Figure 1).

**Figure 1:**
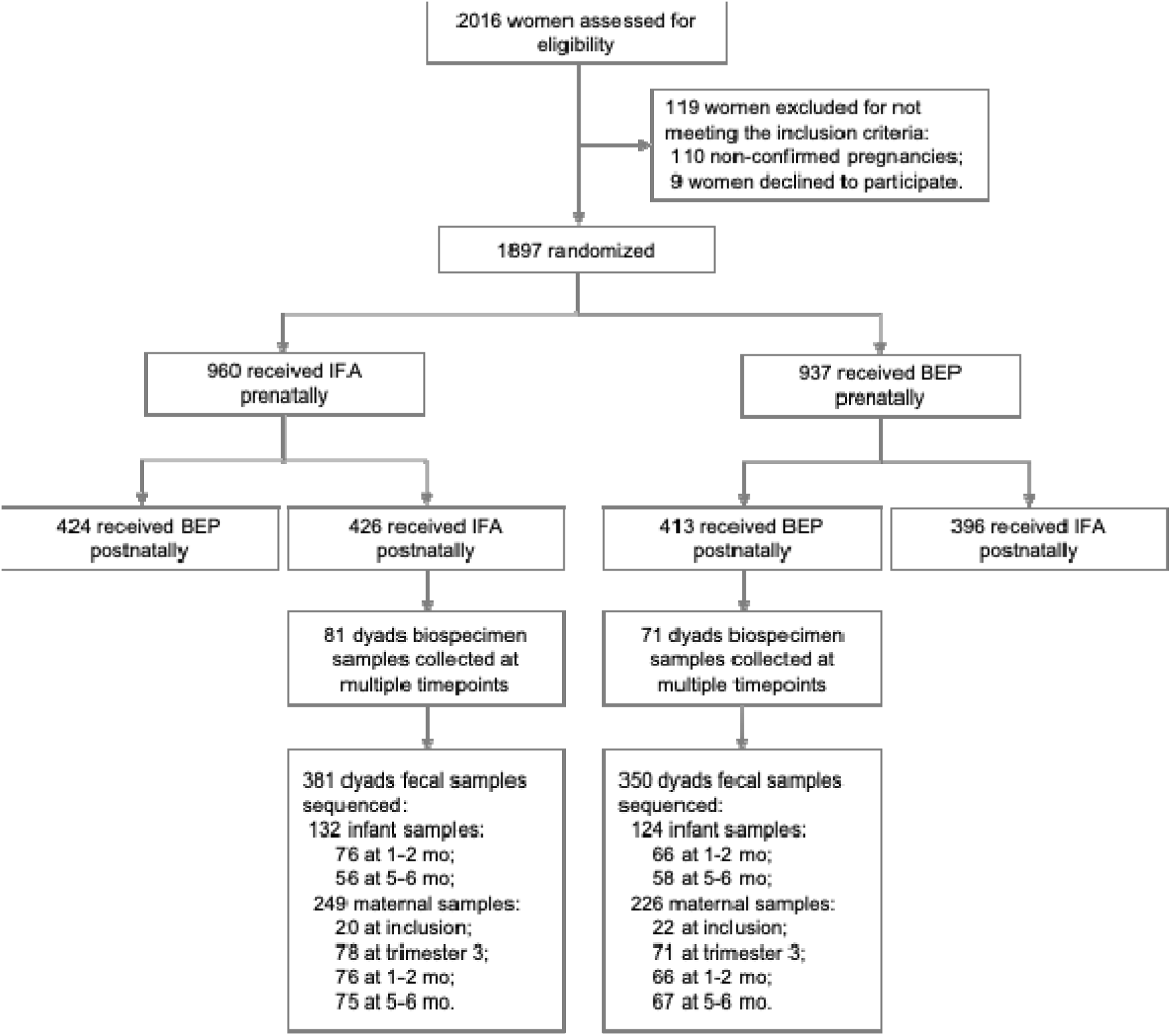
Flow diagram of recruitment, enrollment, and stool sample collection from women and their infants. BEP, balanced energy-protein; IFA, iron-folic acid.

Baseline characteristics of study participants are presented in Table 1. The Mean age of the study women was 24.27 (SD: 5.55) years, and the average body-mass index (BMI) was 22.37 kg/m^2^. The Mean gestational age at birth was 39.82 (SD: 1.72) weeks and more than one in three had at least three prior pregnancies. More than half of the women were from food-insecure households, and 30.3% were anemic. Almost all infants were exclusively breastfed, with an average duration of 5.78 months. Among all participants, the average compliance rate of consuming the BEP supplement was 83.1% during the prenatal and 86.0% during the postnatal supplementation periods.

**Table 1.** Baseline characteristics of study participants.

| Characteristics | Total (n = 152) | BEP group (n = 71) | IFA group (n = 81) |
| --- | --- | --- | --- |
| **Health center catchment area, %** |  |  |  |
| Boni | 14.5 | 15.5 | 13.4 |
| Donhoun | 10.5 | 11.3 | 9.9 |
| Dougoumato II | 18.4 | 21.1 | 16.0 |
| Karaba | 12.5 | 12.7 | 12.3 |
| Kari | 21.7 | 21.1 | 22.2 |
| Koumbia | 18.4 | 15.5 | 21.0 |
| **Household level** |  |  |  |
| Household food insecurity*, % | 59.9 | 62.0 | 58.0 |
| Wealth index, 0 to 10 poits | 4.64 ± 1.75 | 4.69 ± 1.67 | 4.59 ± 1.82 |
| **Maternal** |  |  |  |
| Age, years | 24.27 ± 5.55 | 23.37 ± 5.56 | 25.06 ± 5.46 |
| Ethnic group, % |  |  |  |
| Bwaba | 52.0 | 49.3 | 54.3 |
| Mossi | 35.5 | 39.4 | 32.1 |
| Others | 12.5 | 11.3 | 13.6 |
| Religion of pregnant women, % |  |  |  |
| Animist | 22.4 | 22.5 | 22.2 |
| Muslim | 45.4 | 47.9 | 43.2 |
| Catholic | 6.6 | 7.0 | 6.2 |
| Protestant | 23.0 | 19.7 | 25.9 |
| No religion, no animist | 2.6 | 2.8 | 2.5 |
| Primary education and above, % | 42.1 | 38.0 | 45.7 |
| Number of jobs, % |  |  |  |
| 0 | 58.6 | 64.8 | 53.1 |
| 1 | 38.8 | 33.8 | 43.2 |
| 2 and above | 2.6 | 1.4 | 3.7 |
| Gestational age at birth, weeks | 39.82 ± 1.72 | 40.04 ± 1.63 | 39.63 ± 1.79 |
| Parity, % |  |  |  |
| 0 | 25.0 | 29.6 | 21.0 |
| 1-2 | 36.8 | 33.8 | 39.5 |
| 3 or more | 38.2 | 36.6 | 39.5 |
| Weight, kg | 59.10 ± 10.62 | 57.73 ± 8.30 | 60.30 ± 12.23 |
| Height, cm | 162.35 ± 5.87 | 162.13 ± 4.96 | 162.55 ± 6.59 |
| BMI, kg/m<sup>2</sup> | 22.37 ± 3.47 | 21.97 ± 3.16 | 22.71 ± 3.71 |
| Hb, g/dL | 11.88 ± 1.38 | 11.59 ± 1.47 | 11.73 ± 1.43 |
| Anemia, Hb <11 g/dL, % | 30.3 | 26.8 | 33.3 |
| Number of months receiving EBF | 5.78 ± 1.01 | 5.91 ± 0.74 | 5.67 ± 1.20 |

Values are percentages or means ± SDs.
\*Assessed using FANTA/USAID's Household Food Insecurity Access Scale.
EBF, exclusive breastfeeding; BMI, body mass index; Hb, hemoglobin; SD, standard deviation.

### Deep sequencing of gut microbiome and the early colonization

Mean total sequencing depth of infant and maternal metagenomes was 8.03 (SD: 1.87) and 25.29 (SD: 5.28) giga base-pairs, respectively (Figure S1). 7.4% (SD: 1.7%) and 19.9% (SD: 18.8%) of metagenomic reads were removed from maternal and infant samples during QC and human read decontamination. 58.1% (SD: 14.2%) of infant and 58.0% (SD: 3.0%) maternal reads could be robustly assigned to genomes in our microbial genome database. Out of the 2,654 unique genomes detected in at least one sample, 2,493 are unique to the maternal population, 396 are unique to the infant population, and 235 are observed in both groups. Among the shared microbial genomes, 205 are already observed in the mothers at prenatal time points. The maternal gut microbiomes were dominated by *Prevotella* species, representing a median of 37.9% of the composition, with no major shift between pre- and postnatal compositions (38.1% and 37.7%). In the infant gut microbiomes, *Prevotella* comprised between 0.0% and 83.9% of the gut microbiome, despite being only observed in 44 out of the 256 infant samples. In most infant samples, *Bifidobacterium* strains were dominant, comprising a median of 44.5% of the microbial compositions and present in 98.0% of infant samples (Figure 2A).

**Figure 2:**
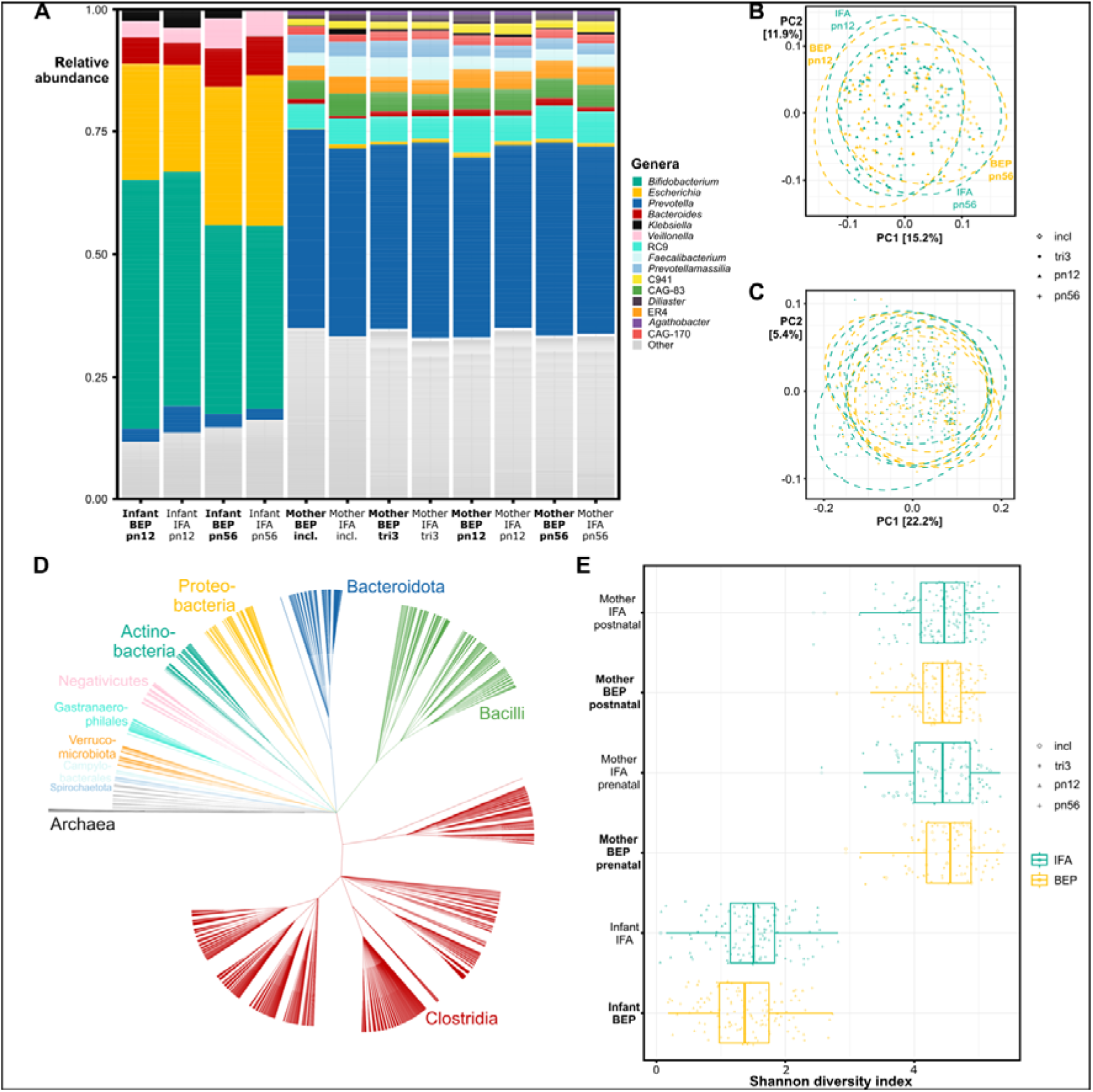
(A) Taxonomic compositions of the top-15 most abundant genera (on average across all samples) aggregated on the treatment- and time groups. The lesser abundant genera are grouped into the “Other” category. **(B)** A PCoA ordination plot of the infant samples. Samples are colored based on the treatment group. **(C)** A PCoA ordination plot of the maternal samples, colored on treatment group. **(D)** The taxonomic tree of the strains identified across all samples, colored on phylum. **(E)** Combined box- and jitter plots of the Shannon diversity indices of the samples, grouped on pre- and postnatal, and on whether the sample was isolated from the infant or the mother, colored on treatment group. Abbreviation: pn12, postnatal 1-2 months; pn56, postnatal 5-6 months; incl., inclusion; tri3, trimester 3.

### BEP supplementation has minimal effect on prenatal gut microbiome composition and diversity

The richness of the infant microbiota is markedly smaller than those of the mothers, with an observed species index between 2 and 43 in the former and between 75 and 614 in the latter. The Shannon diversity index has a median of 1.48 bits in the infant samples and 4.47 bits in the maternal samples and is on average higher in the prenatal samples than in the postnatal ones (Figure 2E). Using Targeted Maximum Likelihood Estimator (TMLE), a significant (*p* = 0.037) effect of BEP is observed on the prenatal maternal microbiome diversity. The average treatment effect is estimated at an increase in Shannon diversity index of 0.144 bits caused by BEP supplementation (95% CI: 0.009 - 0.280). In effective numbers, this translates to an estimated increase of 1.16 equally common species due to the treatment. No significant average treatment effect was found on the postnatal (*p* = 0.97) and infant *(p* = 0.11) gut microbiome diversities.

As illustrated in Figure 2B and 2C, beta-diversity analyses show diversification of the communities, with significant differences between samples taken at different stages of pregnancy (*p* = 0.002) or infancy (*p* = 0.001). In the maternal samples, a marginally significant (*p* = 0.098) inter-community difference is attributed to the BEP supplementation, though this effect is no longer observed in the infant samples (*p* = 0.960).

Differential abundance analyses in the maternal stool samples show significant associations between an *Oscillospiraceae* species [CAG-103 sp 000432375; *q*-value = 2.48×10^-4^; log fold change (logFC) = -1.12] and *Bacteroides fragilis* (*q*-value = 3.26×10^-8^; logFC = 1.31) and BEP supplementation, at inclusion and 5-6 months postpartum, respectively. No robustly significant associations were found between BEP supplementation and the microbial abundances in the infant stool at either time point (Figure 3 and Table S2).

**Figure 3:**
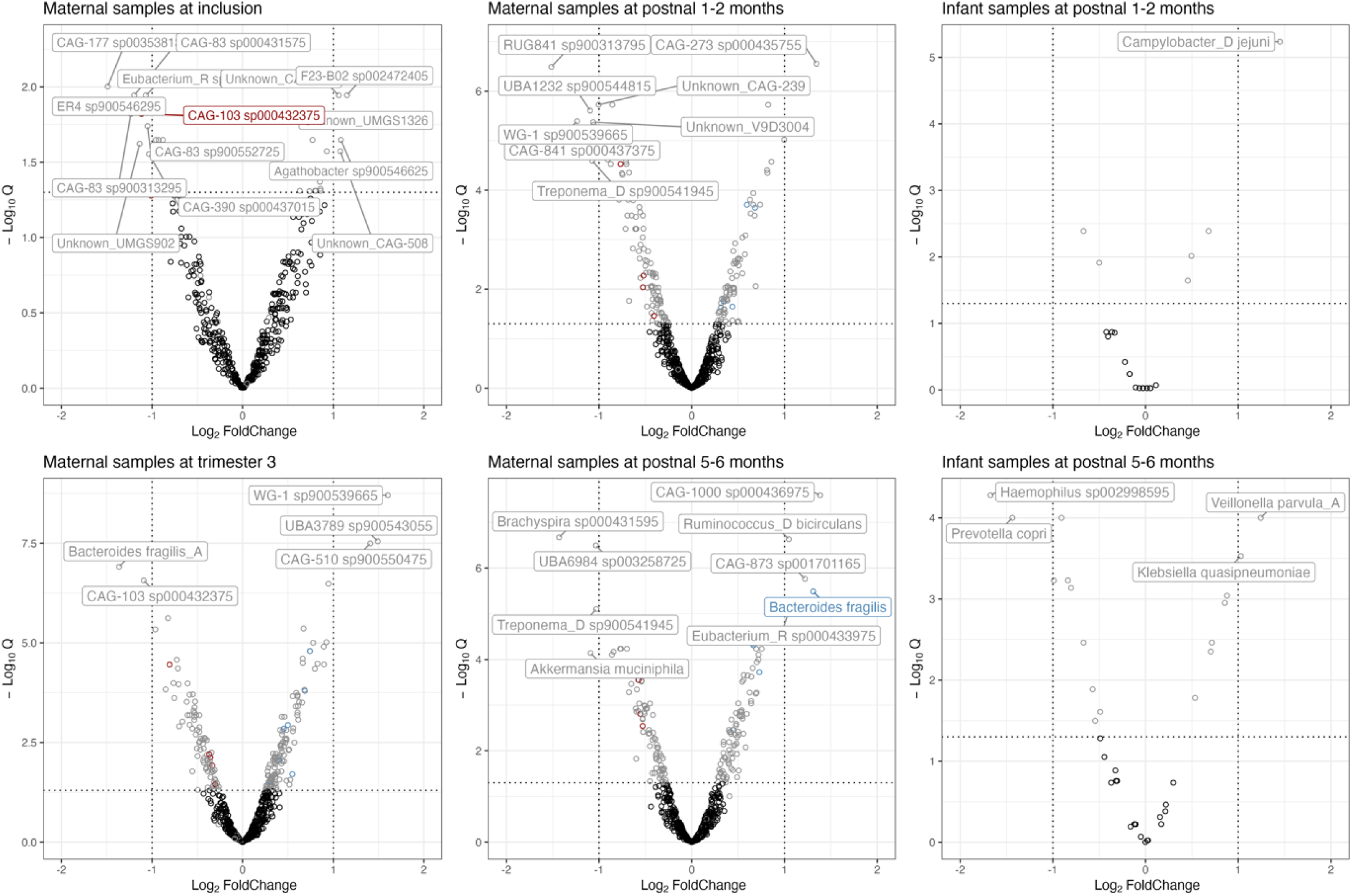
Volcano plots showing differential taxa between two treatment groups at different time points, by ANCOM-BC2. Each point represents an individual taxon, and the position along the x-axis represents logFC in its abundance. The dashed line and labels show the thresholds of statistical and biological significance (Benjamini-Hochberg adjusted *p*-values < 0.05 and |logFC| > 1). The colored dots represent significantly enriched (blue) and depleted (red) taxa that have successfully passed the sensitivity analysis for pseudo-counts. A full table of outputs from ANCOM-BC2 at each time point can be found in Table S2.

### BEP supplementation shows coordinated enrichment of specific microbial pathways

Though no significant association was observed between the treatment intervention and the abundance of individual microbial genes, the Gene Set Enrichment Analysis did indicate concerted enrichment and depletion of certain microbial pathways at each time point (Figure 4). The gene set for the phosphotransferase system is found to be slightly depleted in the stool of BEP-supplemented mothers, both pre- and postpartum (*p*=2.56×10^-5^ at tri3; *p*=2.23×10^-3^ at pn56) and enriched in stool of BEP-supplemented infants (*p*=2.67×10^-2^ at pn12; *p*=2.76×10^-3^ at pn56). Pathways for lipopolysaccharide (LPS) biosynthesis (*p*=1.53×10^-4^ at tri3; *p*=1.88×10^-2^ at pn56), ABC transporters (*p*=2.27×10^-8^ at tri3; *p*=1.17×10^-3^ at pn56), biosynthesis of cofactors (*p*=2.27×10^-8^ at tri3; *p*=1.42×10^-2^ at pn56), and porphyrin metabolism (*p*=2.85×10^-3^ at tri3; *p*=1.74×10^-2^ at pn56) are slightly, but significantly depleted in the BEP supplemented mothers at multiple time points. The pathway for the ribosome complex is significantly enriched in the BEP-supplemented mothers at multiple time points (*p*=1.55×10^-6^ at tri3; *p*=1.17×10^-3^ at pn56). Other pathways, like those for flagellar assembly, are enriched at inclusion (9.35×10^-5^) yet depleted in the third trimester (*p*=1.45×10^-5^) and 1-2 months postpartum (*p*=2.95×10^-8^) in the BEP supplemented mothers. The full table of significantly differential pathways is available in Table S3.

**Figure 4:**
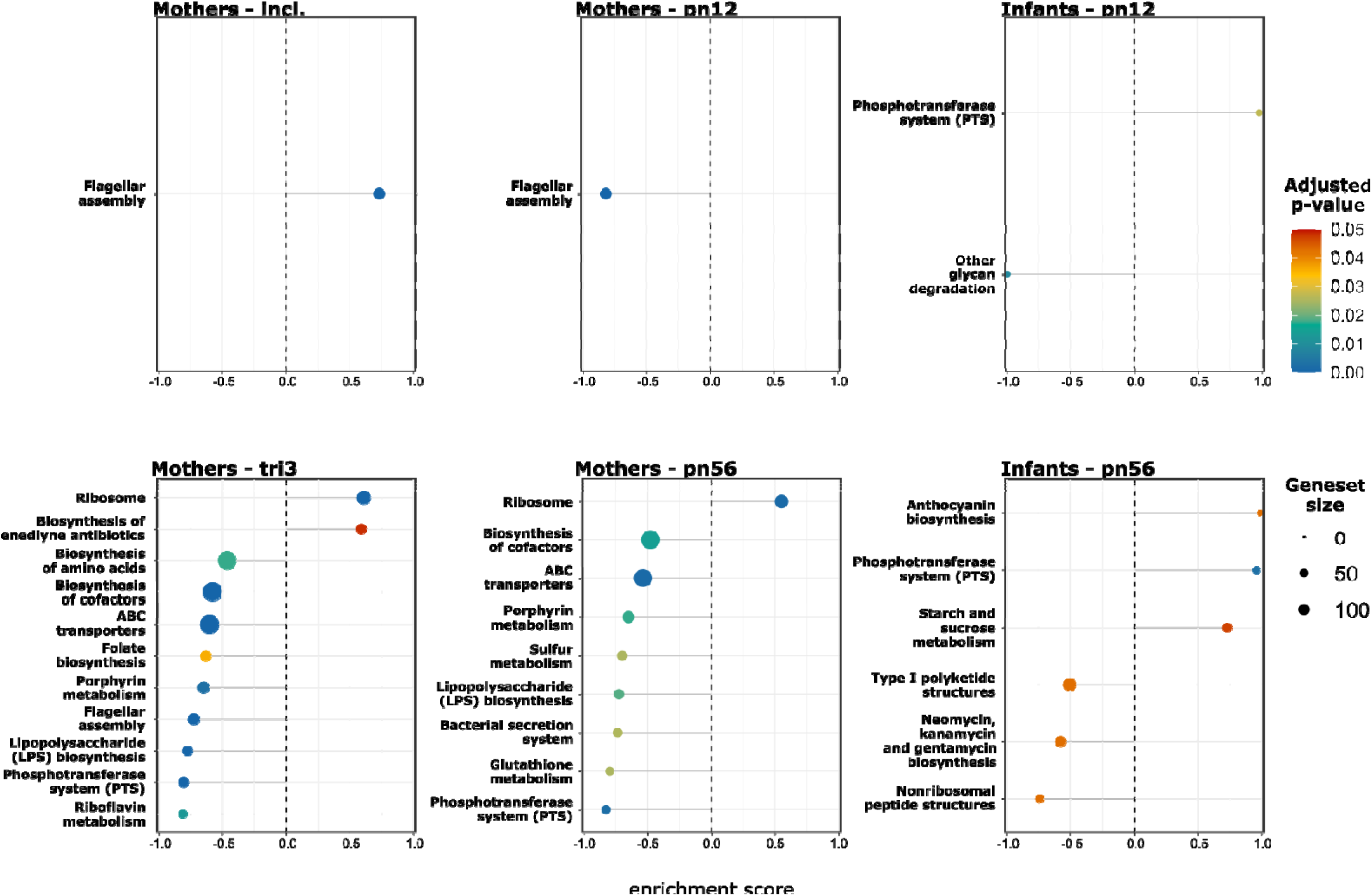
Significantly enriched or depleted KEGG pathways associated to BEP supplementation for each time point. Abbreviation: pn12, postnatal 1-2 months; pn56, postnatal 5-6 months; incl., inclusion; tri3, trimester 3. Points are colored according to adjusted p-value and point size corresponds to gene set size.

In Figure 5, we show that the gene set depletion of the LPS biosynthesis pathway affects specific sections of sequential genes in the pathway. Every gene involved in the conversion from UDP-N-acetyl-D-glucosamine to Lauroyl-KDO2-lipid IV(A) is less abundant with logFC from -0.033 (LpxH) to -0.140 (LpxC). Only one enriched gene, HddA, appears in the LPS biosynthesis pathway in another section of differentially abundant genes following the pentose phosphate pathway. Analogous pathway visualizations are available for the phosphotransferase system, flagellar assembly, and ABC transporters in Supplementary Data (Figure S2-S4).

**Figure 5:**
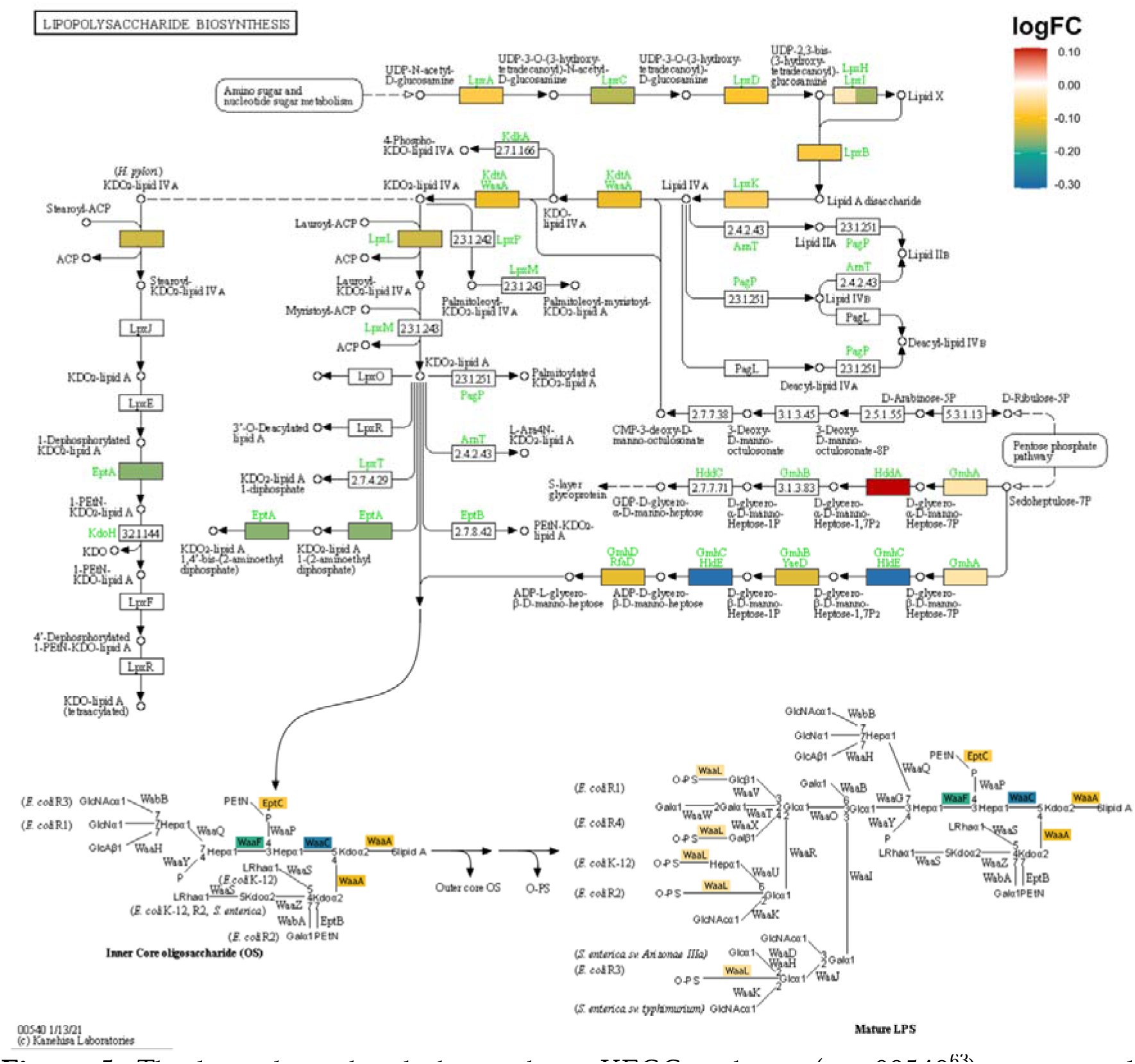
The lipopolysaccharide biosynthesis KEGG pathway (map00540^63^) in maternal samples during the third trimester of pregnancy. Genes colored in yellow, green, and blue are incrementally less abundantly observed in the BEP-supplemented group. Genes colored in red are more abundantly observed in the BEP-supplemented group.

## Discussion

Here, we represent a rare combination of deep sequencing depth and high sample size on a population that is underrepresented in metagenomic literature^25^. We present three novel findings. First, there is a striking disparity in composition of gut microbiome between mothers and their infants in rural Burkina Faso – the infant gut microbiota exhibited a conspicuous dominance of *Bifidobacterium* strains, while *Prevotella* species were dominant in maternal samples. Second, BEP supplementation has minimal effect on maternal gut microbiome – increased Shannon diversity and decreased the abundance of an *Oscillospiraceae* species during pregnancy and increased the abundance of *Bacteroides fragilis* postpartum. Finally, we observed multiple concerted enriched or depleted microbial pathways associated with BEP supplementation, of which, the pathway phosphotransferase system, a major mechanism used by bacteria for uptake of carbohydrates^26^, is enriched in infant samples whose mother were BEP-supplemented.

*Bifidobacterium,* commonly found in exclusively breastfed infants, are crucial in the infant gut because they breakdown human milk oligosaccharides (HMO) to short chain fatty acids to maintain gut health^27,28^. In this study, nearly all infants, regardless of intervention groups, experienced exclusive breastfeeding during the sample collection period. This circumstance might explain the comparability in abundance patterns between the groups and suggests that the provision of BEP during pregnancy and lactation exerted no discernible impact on the primary beneficial colonizers or of their HMO utilization genes in infants. Previous researches highlighting the prevalence of *Prevotella*-rich gut in non-industrialized populations, particularly those adhering to high-fiber diets, in stark contrast to the lower occurrence observed in industrialized and westernized nations^11,17,29^. This distinct enterotype characterized by *Prevotella* enrichment had also been noted in early childhood, spanning a wide age from 7 to 37 months, as observed in a Gambian cohort^29^. Interestingly, we observed a strong tie of *Prevotella* between microbial genomes in mother and infant samples. In the latter, *Prevotella* was either markedly absent or notably abundant, implying its swift dominance within the ecological niche upon establishing residence within the infantile gut.

Gut microbiome alpha diversity has been linked to human health, with lower levels of diversity associated with several acute and chronic disease^30^. Recent studies have reported a noteworthy association between a high-fat diet and a decrease alpha diversity, concomitant with the onset of low-grade inflammation^31,32^. This in turn facilitate the increase and translocation of bacterial products such as pro-inflammatory LPS^33^. Intriguingly, our findings indicate the contrary: an augmentation in alpha diversity and a concurrent decrease in LPS synthesis within the prenatal gut microbiota in women who received BEP. The subsection of the LPS biosynthesis pathway which is depleted, *i.e.*, the central KDO and Lipid A metabolism, is highly conserved and invariant in the gut microbiome of healthy individuals^34^. Given that different covalent modifications of Lipid A also have different immunological properties^35,36^, follow-up research integrating proteomics and metabolomics data is currently ongoing to fully understand the clinical implications of these findings.

The observed significant variation in alpha diversity within the maternal gut microbiota during pregnancy, without a corresponding effect among infants, suggests that the maternal component of BEP supplementation did not exert a detectable influence on the diversity of child’s microbiome. This aligns with the observations stemming from an LNS intervention conducted in rural Malawi^21^.

We observed a significant association between BEP supplementation during the second trimester and a reduction in the abundance of CAG-103 sp000432375, an *Oscillospiraceae* species that has recently been recognized as responsive to carbohydrates^37^. The overall lesser dependence on carbohydrate metabolism can also be observed in the concerted depletion of genes for the phosphotransferase system and ABC transporters in the maternal metagenomes^38,39^. In addition, BEP supplementation is associated with increased abundance of *Bacteroides fragilis* within the maternal gut at 5-6 months postpartum. *B. fragilis* is a major component of the fecal microbiome, with potentially billions of *B. fragilis* within the gastrointestinal tract^40^, and plays a crucial role in the degradation of host-derived mucosal glycans, including mucins and the *N*-link oligosaccharides of glycoproteins^41^. Nevertheless, during the third trimester, a contrasting association was observed between BEP supplementation and another strain of *B. fragilis*. However, it is important to exercise caution when interpreting this finding, as it did not withstand the sensitivity analysis for pseudo-counts, suggesting the possibility of a false positive result.

Despite the potential for false positives, BEP supplementation appears to exhibit an association with the increases in the abundance of *Campylobacter jejuni,* and *Veillonella parvula* and *Klebsiella quasipneumoniae* during the first two months and at the five-to-six months of early life, respectively. In LMICs, infection with *Campylobacter* is one of the major causes of diarrheal disease in children^42^, leading to early protein malnutrition^43^. A recent study has reported higher rates of campylobacteriosis among exclusively breastfed infants in LMICs. It suggests that specific *C. jejuni* strains can utilize HMO and proteins in human milk to promote the growth of campylobacters^44^. Similarly, another study revealed that the content of the secretory immunoglobulin A (sIgA) in breastmilk was positively correlated with the abundance of *V. parvula* in infant gut microbiota, and suggested that this regulation during early life is one of the potential mechanisms by which sIgA in breast milk modulates immune function^45^. *Klebsiella pneumoniae*, comprising *K. quasipneumoniae*, is a leading pathogen for neonatal infection^46,47^. A recent study has reported the association of the high abundance of *Klebsiella* and high content of breast milk N-acetylneuraminic acid (Neu5Ac), the predominant form of free salic acid and important component binding to sialylated oligosaccharides in breast milk, which indicates that *Klebsiella* could utilize sialic acid as carbon source^48^. Hence, the observed connection between BEP supplementation and increased abundance of these species in the infant gut microbiome imply that BEP supplementation during pregnancy and lactation may influence the infant microbiome by impacting the composition of human milk. However, further analyses aimed at evaluating the effect of BEP supplementation on breastmilk composition are imperative to gain a more comprehensive understanding of these observations.

We identified that the phosphotransferase system, a critical mechanism for bacterial carbohydrates uptake, exhibited enrichment in infants born to BEP-supplemented mothers. This implies that the provision of BEP supplementation to mothers may elevate carbohydrate content in human milk, thereby augmenting the bacterial carbohydrates uptake capacity within the infant gut, potentially contributing to enhanced linear growth. However, subsequent analyses, incorporating a detailed evaluation of human milk components and employing a multi-omics approach are imperative for a comprehensive understanding of the intricate biological pathways.

Some significantly differential pathways were found that are unlikely to be of microbial origin, such as those for the ribosome complex, the biosynthesis of cofactors, and porphyrin metabolism. Though the observation of these pathways is likely a limitation of the KEGG database these genes were annotated with, their consistent association to the BEP supplemented metagenome can be indication that there are so far unknown microbial processes that involve these genes.

## Methods

### Study setting and design

The MISAME-III study protocol^49^ and the biospecimen sub-study (Biospé)^50^ were published earlier. Briefly, the MISAME-III trial (NCT03533712) is an individually randomized 2 x 2 factorial RCT evaluating the efficacy of multiple micronutrient-fortified BEP supplementation during pregnancy and lactation on birth outcomes and infant growth in rural Burkina Faso. Biospé study, which was nested within the MISAME-III trial, aimed to evaluate the physiological effects of BEP supplementation on both pregnant and lactating women and their infants.

The study was implemented in six rural health center catchment areas in the district of Houndé in the Hauts-Bassins region of Burkina Faso from October 2019 to December 2020. A total of 1,897 women aged between 15 and 40 years with a gestational aged < 21 completed weeks, and living in the study catchment villages, were enrolled in this trial. The habitual diet during pregnancy is nondiverse^51^, being predominantly based on maize with a supplement of leafy vegetables^52^.

Women were randomly assigned to the prenatal intervention arms receiving either fortified BEP supplements and iron-folic acid (IFA) tablets (i.e., intervention) or IFA alone (i.e., control), which is the standard of care during pregnancy in Burkina Faso. The same women were concurrently randomized to receive either of the postnatal interventions, which comprised fortified BEP supplementation during the first six months postpartum in combination with IFA for the first 6 weeks (i.e., intervention), or the postnatal control, which comprised IFA alone for six weeks postpartum (i.e., control). Therefore, there were four study groups in MISAME-III study: (1) prenatal BEP and IFA supplementation; (2) postnatal BEP and IFA supplementation; (3) both pre- and postnatal BEP and IFA supplementation; or (4) both pre- and postnatal IFA only supplementation.

### Study supplements

The BEP supplement is a lipid-based nutrient supplement in the form of an energy-dense peanut paste fortified with multiple micronutrients. The BEP is produced by Nutriset and is ready-to-consume, does not require a cold chain and is highly stable with a long shelf life. The complete nutritional composition of a daily dose of the fortified BEP is provided in S1 Table. On average, a daily dose of the BEP (72g) provided an energy top up of 393 kcal (36% energy from lipids, 20% from protein, and 32% from carbohydrates) and covered at least the estimated average nutritional requirements of pregnant women for 11 micronutrients, except calcium, phosphorous, and magnesium^53^. A daily dose of an IFA tablets (Sidhaant Life Science, Delhi, India) contained 65 mg of iron [form: FeH_2_O_5_S] and 400 µg folic acid [form: C_19_H_19_N_7_O_6_]. Following Burkinabè guidelines, all enrolled women received malaria prophylaxis (three oral doses of sulfadoxine-pyrimethamine).

### Fecal sample collection

As a sub-analysis of Biospé study, fecal samples from mother-infant dyads were collected in all four groups but only two groups of samples, both pre- and postnatal BEP and IFA supplementation group (BEP group), and both pre- and postnatal IFA only supplementation group (IFA group) have been sequenced. Maternal fecal samples (8g) were collected in a fecal pot and then aliquoted into sterile cryotubes (Biosigma, Cona, VE, Italy), flash-frozen and transferred to a -80 ◦C freezer. Infant feces (8g) were collected using a 38 × 50 cm sterile protection sheet (Kimberley-Clark, Irving, TX, USA) which is used like a diaper and wrapped around the newborn, before they were transferred to a OMNIgene•GUT OM-200 collection kit and then sterile cryotubes for storage at -80 ◦C. The collected feces were assessed for consistency based on the visual Bristol scale. For liquid feces, thorough homogenization was performed using a plastic spoon to mix the solid and liquid.

Fecal samples were collected from the pregnant and lactating women at inclusion into the Biospé study when most of the women were in trimester 2 (19-24 weeks) and have been taken supplementation for 2-3 weeks, trimester 3 (30-34 weeks) and 1-2 (28-56 days) and 5-6 months (147-175 days) postpartum. Infant fecal samples were collected in 1-2 months (12-56 days of life) and 5-6 months (140-168 days of life) corresponding to maternal postnatal time points.

### Metagenomic library preparation and sequencing

DNA extraction, library preparation, and metagenomic sequencing was performed at the UC San Diego IGM Genomics Center. DNA extraction was performed using the Thermo MagMAX Microbiome Ultra kit. DNA quantification was performed using picogreen dye. Roche KAPA HyperPlus and 96-UDI plates were used for library preparation. In this study, 25ng of input DNA and 9 PCR cycles were used during library preparation. A shallow MiSeq Nano sequencing run was used to normalize read sequencing depth. 2 x 150bp reads were then generated on the Illumina NovaSeq 6000 at a target depth of 25 giga base-pairs per sample for maternal samples and 10 giga base-pairs per sample for infant samples.

### Metagenomic read annotation

The metagenomic sequencing reads were trimmed and deduplicated using FastP. All reads mapping to the hg38 human genome with Bowtie2 under default settings were removed. Samples were profiled by mapping preprocessed reads from each sample to a previously established database of microbiome genomes (database “DeltaI”) using Bowtie2 with default settings. The DeltaI microbial genome database was specifically created to profile non-industrialized infant microbiomes^13^. The resulting mapping files were analyzed using inStrain profile under default settings^54^. Relative abundance of each genome was calculated using the formula: number of reads mapping to genome / number of processed reads in sample. Only genomes with breadth > 0.5 were considered in analyses.

Gene-level functional profiling was performed using the inStrain “parse_annotations” pipeline with default settings (https://instrain.readthedocs.io/en/master/user_manual.html#parse-annotations). Genes were called for each genome individually using Prodigal in “single” mode, and all gene files were then concatenated together. Functional annotation was performed against (1) Kyoto Encyclopedia of Genes and Genomes (KEGG) Orthologies (KOs) using kofam_scan (version 1.3); (2) carbohydrate-active enzymes (CAZymes) using dbCAN2 (version 11); (3) Comprehensive Antibiotic Resistance Database (CARD) Anti-Microbial Resistance (AMR) database version 3.2.5 using Diamond with the command “diamond blastp -f 6 -e 0.0001 -k 1”; and (4) HMO proteins using the instructions provided here: https://instrain.readthedocs.io/en/master/user_manual.html#human-milk-oligosaccharide-hmo-utilization-genes.

### Taxonomic data analysis

Alpha-diversity metrics for Observed Species index, Simpson’s Diversity index, and Shannon Diversity index were calculated in R using phyloseq v1.44.0^55^. The treatment effect of the randomized intervention was estimated by the doubly robust TMLE in R using tmle v1.5.0^56^. Given the likely heterogeneity of the effect between the populations, this estimation was performed for maternal and infant data separately, further separated into pre- and postnatal for the maternal samples. Maternal stratum data (level of education, marital status, language, religion, ethnicity, number of jobs, socio-economic status, dietary diversity score, and health center), as well as anthropometric data taken at inclusion into the trial (age, height, BMI and hemoglobin level) were taken as covariates for the model. Both covariate-outcome and propensity score models were fitted with ensemble SuperLearner (v2.0-28) models. Diversity indices were translated to effective numbers of species using the formulas posed by Jost^57^.

Beta-diversity analysis was performed in R using vegan v2.6-4. Ordination was performed on the weighted UniFrac distances between the log transformed strain counts, followed by multivariate statistics between groups using permutational multivariate analysis of variance (PERMANOVA) with default parameters.

To address compositionality and zero-inflation issues of microbial count data, the differential abundance analyses was performed by using Analysis of Compositions of Microbiomes with Bias Correction (ANCOM-BC) v2.2.1 at every taxonomic level separately to identify taxa associated to the treatment intervention^58^. Benjamini-Hochberg multiple testing correction was used, and the alpha-level was set at 0.05. To avoid inflated false positive rate, a sensitivity analysis was conducted against the addition of pseudo-counts. Only robustly significant taxa were reported.

### Functional data analysis

A hierarchical differential abundance analyses was conducted on the Trimmed Mean of M-values (TMM) library sizes for the gene counts using treeclimbR v0.1.5 and edgeR in R^59^. Genes from KEGG, HMO, CAZyme and CARD were categorized into a hierarchical structure based on their respective Enzymatic Class (EC) number, functional group, HMO cluster or antibiotic resistance mechanism. Benjamini-Hochberg multiple testing correction was used, and the alpha-level was set at 0.05.

Using the KEGG pathway mappings, we performed a Gene Set Enrichment Analysis to identify pathways that were differentially abundant in a coordinated direction^60^. A *Bifidobacterium longum* NagR regulon gene set was also added for the HMO utilization genes found in Table 1 of Arzamasov et. al (2022)^61^. For each set, an enrichment score with a corresponding adjusted p-value was calculated in R using clusterProfiler v4.2.2^62^. The alpha-level was set at 0.05 and the minimum gene set size to 1.

### Ethics

The study was approved by the Commissie voor Medische Ethiek (CME) of Ghent University Hospital (protocol code: B670201734334 and date of 10/08/2020) and the Comité d’Éthique Institutionnel de la Recherche En Sciences de la Santé (CEIRES) of the Institut de Recherche en Sciences de la Santé (IRSS) (protocol code 50-2020/CEIRES and date of 22/10/2020). An independent Data and Safety Monitoring Board (DSMB), comprising an endocrinologist, two pediatricians, a gynecologist, and an ethicist of both Belgian and Burkinabe nationalities, was established prior to the start of the efficacy trial. The DSMB managed remote safety reviews for adverse and serious events at nine and 20 months after the start of enrolment. The MISAME-III trial was registered on *ClinicalTrials.gov* (identifier: NCT03533712).

## Author Contributions

LD and ST performed the statistical analyses, visualized data, interpreted data, and prepared the manuscript with contributions from all authors. MRO, EDS, and JLS were responsible for the sample analysis, interpreted preliminary data and critically reviewed the manuscript. TDC, LCT, CL and PK conceived of the study, acquired funding, and critically reviewed the manuscript. TDC, LCT, LO, YBM, MO, RG, CL, and PK set up the study and supervised the project. TDC, LO, YBM, MO and RG collected samples. TDC, EB, AA, KT, WVC, and MS supervised the data curation and contributed to data interpretation. All authors have read and agreed to the published version of the manuscript.

## Data Availability

Given the personal nature of the data, data will be made available through a data-sharing agreement. Please contact for any queries. Supporting study documents, including the study protocol and questionnaires, are publicly available on the studys website: https://misame3.ugent.be (accessed on 23 November 2023).

## Acknowledgements

The work is supported by the Bill & Melinda Gates Foundation (OPP1175213). Lishi Deng is supported by the China Scholarship Council (Grant No. 202207650056). Steff Taelman is supported by the Flemish Agency for Innovation and Entrepreneurship (VLAIO HBC.2020.2292). This publication includes data generated at the UC San Diego IGM Genomics Center utilizing an Illumina NovaSeq 6000 that was purchased with funding from a National Institutes of Health SIG grant (#S10 OD026929). The funders had no role in the design and conduct of the study; in the collection, management, analysis, and interpretation of the data; or in the preparation, review, or approval of the manuscript.

The authors thank all participants from Burkina Faso and the data collection team. We thank Nutriset (France) for donating the BEP supplements.

## Data Availability Statement

The informed consent form does not allow sharing of personal data outside the research team. Data will be made available through a data-sharing agreement. Please contact for any queries. Supporting study documents, including the study protocol and questionnaires, are publicly available on the study’s website: https://misame3.ugent.be (accessed on 23 November 2023).

## Informed Consent Statement

Informed consent was obtained from all subjects involved in the study.

## Competing Interests

The authors have declared no competing interests.

**Table S1**: Nutritional values of the ready-to-use supplementary food for pregnant and lactating women.

**Table S2**: Results of the differential abundance analyses between intervention groups.

**Table S3**: Results of the gene set enrichment analyses between intervention groups.

**Figure S1:**
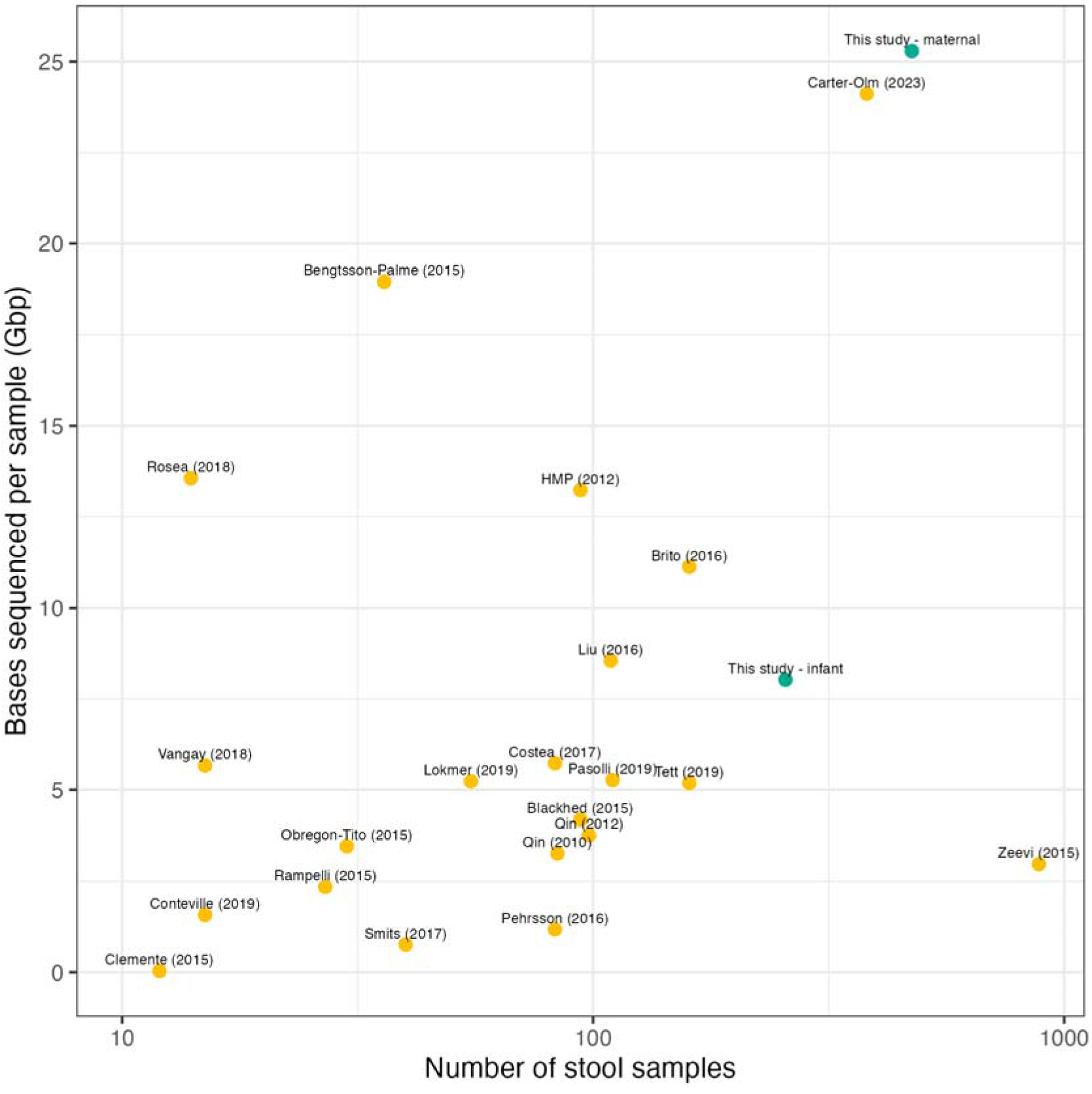
The number of samples versus the number of bases sequenced per sample for various gut metagenomic datasets and this study (adapted from (Carter et al., 2023)^64^).

**Figure S2:**
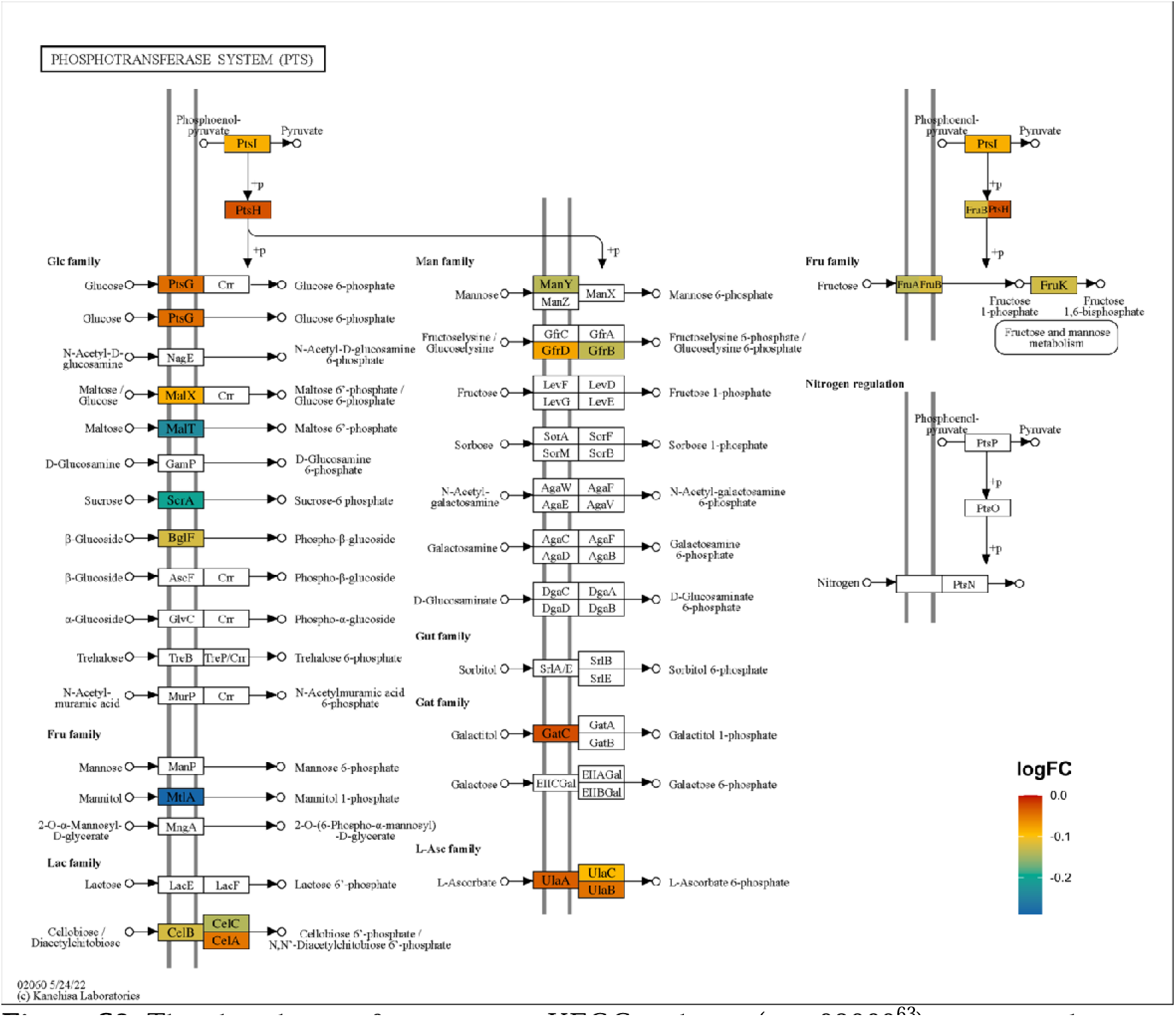
The phosphotransferase system KEGG pathway (map02060^63^) in maternal samples during the third trimester of pregnancy. Genes colored in red, yellow, green, and blue are incrementally less abundantly observed in the BEP-supplemented group.

**Figure S3:**
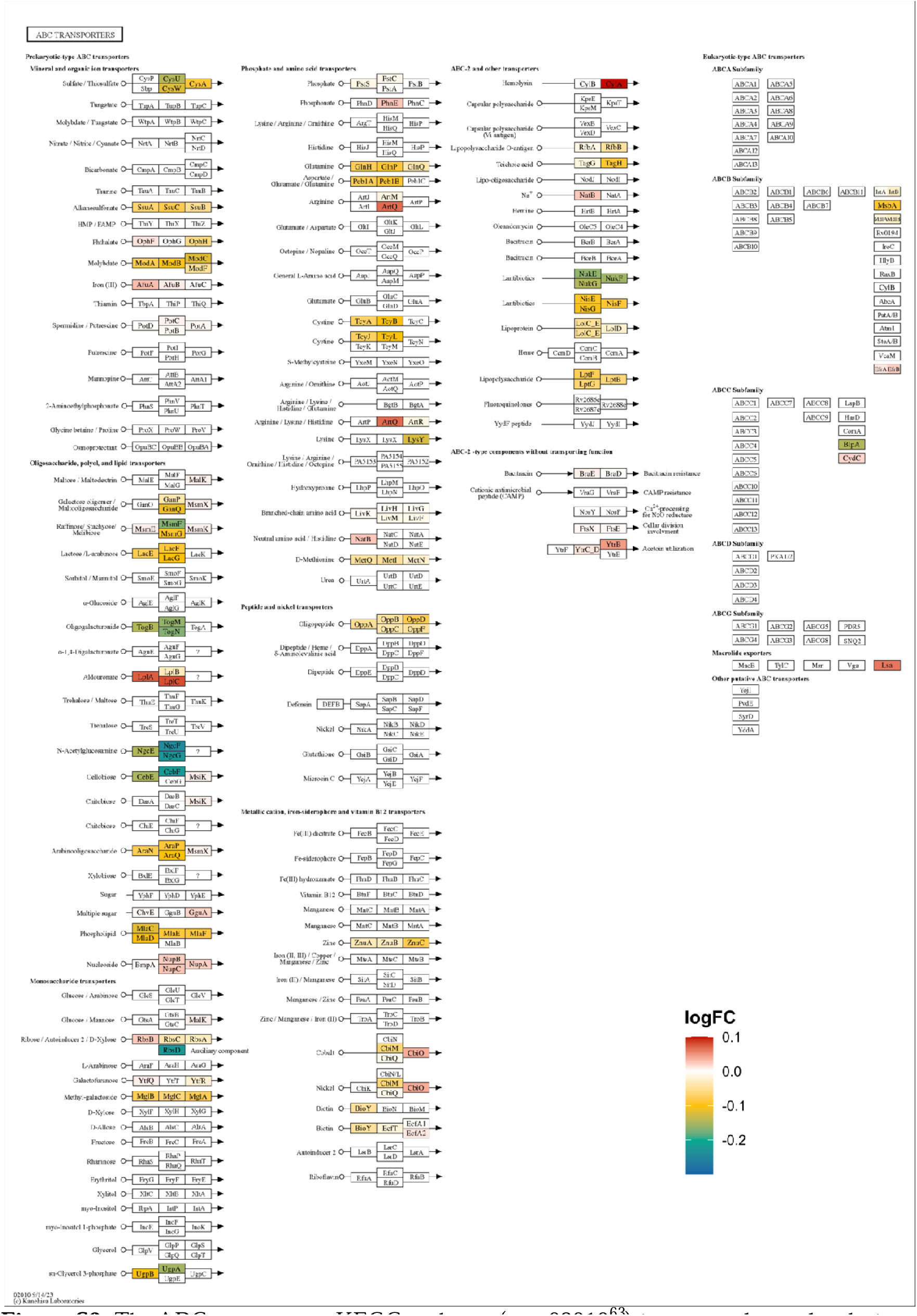
The ABC transporters KEGG pathway (map02010^63^) in maternal samples during the third trimester of pregnancy. Genes colored in yellow, green, and blue are incrementally less abundantly observed in the BEP-supplemented group. Genes colored in red are more abundantly observed in the BEP-supplemented group.

**Figure S4:**
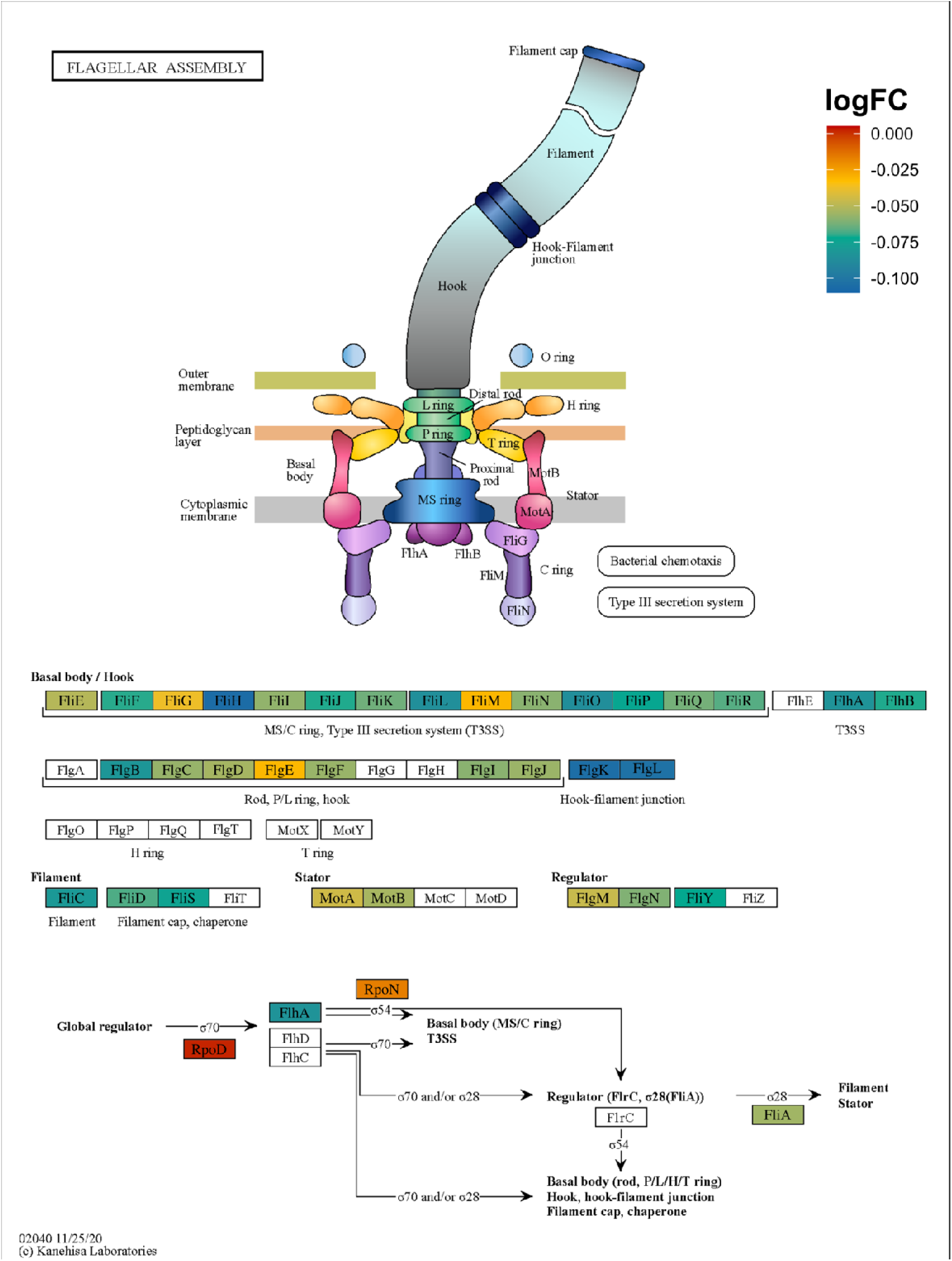
The flagellar assembly KEGG pathway (map02040^63^) in maternal samples during the third trimester of pregnancy. Genes colored in red, yellow, green, and blue are incrementally less abundantly observed in the BEP-supplemented group.

---

CONSORT 2010 checklist of information to include when reporting a randomised trial*.

| Section/Topic | Item No | Checklist item | Reported on page No |
| --- | --- | --- | --- |
| **Title and abstract** |  |  |  |
|  | 1a | Identification as a randomised trial in the title | 1 |
|  | 1b | Structured summary of trial design, methods, results, and conclusions (for specific guidance see CONSORT for abstracts) | 1 |
| **Introduction** |  |  |  |
| Background and objectives | 2a | Scientific background and explanation of rationale | 2, 3 |
|  | 2b | Specific objectives or hypotheses | 3 |
| **Methods** |  |  |  |
| Trial design | 3a | Description of trial design (such as parallel, factorial) including allocation ratio | 9 |
|  | 3b | Important changes to methods after trial commencement (such as eligibility criteria), with reasons | 9 |
| Participants | 4a | Eligibility criteria for participants | 9 |
|  | 4b | Settings and locations where the data were collected | 9 |
| Interventions | 5 | The interventions for each group with sufficient details to allow replication, including how and when they were actually administered | 9, 10 |
| Outcomes | 6a | Completely defined pre-specified primary and secondary outcome measures, including how and when they were assessed | 9, 10 |
|  | 6b | Any changes to trial outcomes after the trial commenced, with reasons | NA |
| Sample size | 7a | How sample size was determined | 3 |
|  | 7b | When applicable, explanation of any interim analyses and stopping guidelines | NA |
| **Randomisation:** |  |  |  |
| Sequence generation | 8a | Method used to generate the random allocation sequence | 9 |
|  | 8b | Type of randomisation; details of any restriction (such as blocking and block size) | 9 |
| Allocation concealment mechanism | 9 | Mechanism used to implement the random allocation sequence (such as sequentially numbered containers), describing any steps taken to conceal the sequence until interventions were assigned | 9 |
| Implementation | 10 | Who generated the random allocation sequence, who enrolled participants, and who assigned participants to interventions | NA |
| Blinding | 11a | If done, who was blinded after assignment to interventions (for example, participants, care providers, those assessing outcomes) and how | NA |
|  | 11b | If relevant, description of the similarity of interventions | NA |
| Statistical methods | 12a | Statistical methods used to compare groups for primary and secondary outcomes | 10 - 12 |
|  | 12b | Methods for additional analyses, such as subgroup analyses and adjusted analyses | NA |
| **Results** |  |  |  |
| Participant flow (a diagram is strongly recommended) | 13a | For each group, the numbers of participants who were randomly assigned, received intended treatment, and were analysed for the primary outcome | 3 |
|  | 13b | For each group, losses and exclusions after randomisation, together with reasons | 3 |
| Recruitment | 14a | Dates defining the periods of recruitment and follow-up | 9 |
|  | 14b | Why the trial ended or was stopped | NA |
| Baseline data | 15 | A table showing baseline demographic and clinical characteristics for each group | 15 |
| Numbers analysed | 16 | For each group, number of participants (denominator) included in each analysis and whether the analysis was by original assigned groups | 3 |
| Outcomes and estimation | 17a | For each primary and secondary outcome, results for each group, and the estimated effect size and its precision (such as 95% confidence interval) | 3 - 5 |
|  | 17b | For binary outcomes, presentation of both absolute and relative effect sizes is recommended | NA |
| Ancillary analyses | 18 | Results of any other analyses performed, including subgroup analyses and adjusted analyses, distinguishing pre-specified from exploratory | NA |
| Harms | 19 | All important harms or unintended effects in each group (for specific guidance see CONSORT for harms) | NA |
| **Discussion** |  |  |  |
| Limitations | 20 | Trial limitations, addressing sources of potential bias, imprecision, and, if relevant, multiplicity of analyses | 8 |
| Generalisability | 21 | Generalisability (external validity, applicability) of the trial findings | 5, 6 |
| Interpretation | 22 | Interpretation consistent with results, balancing benefits and harms, and considering other relevant evidence | 6 - 8 |
| **Other information** |  |  |  |
| Registration | 23 | Registration number and name of trial registry | 13 |
| Protocol | 24 | Where the full trial protocol can be accessed, if available | 14 |
| Funding | 25 | Sources of funding and other support (such as supply of drugs), role of funders | 13 |

Citation: Schulz KF, Altman DG, Moher D, for the CONSORT Group. CONSORT 2010 Statement: updated guidelines for reporting parallel group randomised trials. BMC Medicine. 2010;8:18. © 2010 Schulz et al. This is an Open Access article distributed under the terms of the Creative Commons Attribution License (http://creativecommons.org/licenses/by/2.0), which permits unrestricted use, distribution, and reproduction in any medium, provided the original work is properly cited.
\*We strongly recommend reading this statement in conjunction with the CONSORT 2010 Explanation and Elaboration for important clarifications on all the items. If relevant, we also recommend reading CONSORT extensions for cluster randomised trials, non-inferiority and equivalence trials, non-pharmacological treatments, herbal interventions, and pragmatic trials. Additional extensions are forthcoming: for those and for up-to-date references relevant to this checklist, see www.consort-statement.org.

